# Assessment of the accuracy of lung lesions diagnosis in adolescents with osteosarcoma using artificial intelligence

**DOI:** 10.64898/2026.06.08.26354011

**Authors:** Natalya G. Uskova, Victor A. Gombolevskiy, Valeriya Yu. Chernina, Dmitriy V. Burenchev, Dmitriy G. Akhaladze, Elena V. Panina, Aleksandr I. Karachunskiy, Galina V. Tereschenko, Mikhail Yu. Goncharov, Evgeniya A. Soboleva, Elena I. Konopleva, Oleg I. Bydanov, Sergey Yu. Plekhov, Nikolay S. Grachev

## Abstract

**Background:** Lung metastases in osteosarcoma (OS) are the main cause of the death. The accuracy of the diagnosis of nodules by computed tomography (CT) of the lungs is critically important for determining the disseminated stage of the disease and planning surgical treatment. The use of artificial intelligence (AI) in the search for lung nodules increases the accuracy of diagnosis and reduces the chance of missing metastases.

**Objective:** to evaluate the accuracy of lung nodules diagnosis in adolescents with OS using AI.

**Methods:** A retrospective assessment of CT scans of adolescents with OS was performed. A pathological nodule with an average size of ≥4 mm was considered a target finding. The diagnostic accuracy of an AI algorithm previously trained on an adult dataset was evaluated, and the number of false positives (FP) and false negatives (FN) was determined. Sensitivity, specificity, accuracy, area under the ROC curve (AUC), positive predictive value, negative predictive value, and F1- measure were calculated. Based on the obtained results, the effectiveness of the algorithm was assessed.

**Results:** 248 CT scans of adolescents with OS were evaluated. The following results were obtained: in 5 cases, the AI algorithm showed a FP result (2%), in 34 cases, it showed a FN result (13.7%), and in 209 cases, a correct result (both true positive and true negative) (84.3%). The diagnostic accuracy of the algorithm was 0.843 (95% CI 0.794-0.887). The application of the AI algorithm in the practice of an X-ray doctor in a specific clinical task would allow to increase the sensitivity from 0.805 to 0.891, while ensuring an absolute decrease in the number of FN results by 8.6% and a relative decrease by 44%.

**Conclusion:** The obtained results confirm the practical value of the application of the AI algorithm and justify the implementation of AI-assisted systems in the diagnostic protocols for lung metastases in adolescents with OS.

## BACKGROUND

Osteosarcoma (OS) is the most common primary bone sarcoma in adolescents and young adults [1]. At the time of diagnosis, pulmonary metastases are detected in 20% of patients [2], which constitutes an adverse prognostic factor and reduces 5-year overall survival from 85% to 57% (unpublished institutional data); according to Rasalkar et al., survival decreases from 60-70% to 10-30% [3]. These data indicate that accurate detection of pulmonary lesions on chest computed tomography (CT) is critical for timely assignment of disseminated disease stage and for optimizing treatment of metastases [4]. It is well established that human factors, limited workforce capacity, and the high workload of staff in the radiology department of a referral center may lead to missed pathological findings on CT, inevitably resulting in negative clinical consequences for patients [5, 6].

Artificial intelligence (AI) based on computer vision can analyze chest CT images, detect pathological findings (in OS, pulmonary lesions, including calcifications), assess their linear dimensions and volume, mark them, and generate a text description and interpretation of the study [4, 7, 8, 9, 10, 11, 12].

In contemporary oncology, AI is actively used for segmentation of magnetic resonance images of affected bones in skeletal malignant neoplasms (MN) [13, 14, 15, 16]. However, the number of publications on the diagnosis of pulmonary metastases of MN is very limited, especially in pediatric patients [10, 11, 12]; moreover, the study by Ni et al. is the only one indexed in PubMed that is dedicated to the diagnosis of OS metastases in children and adolescents [10].

### Study objective

to evaluate the diagnostic accuracy of pulmonary lesion detection on chest CT using an AI algorithm in adolescents with OS.

## METHODS

### Study design

a retrospective multicenter cross-sectional clinical study.

### Study setting

The study included data from patients who received any type of medical care – consultative, therapeutic, surgical, combined, or comprehensive – at the Dmitry Rogachev National medical research center of pediatric hematology, oncology and immunology (Dmitry Rogachev NMRC PHOI), Moscow, Russia (hereinafter, the Center) between January 2012 and March 2025. Patients underwent initial evaluation, including chest CT, not only at the Center but also in various regions of the Russian Federation.

The dataset comprised CT examinations from patients enrolled in the Center’s Scientific Registry of Bone and Soft Tissue Tumors, which has been maintained since January 2012 and includes database registration of patients with MN of bone and soft tissues. The database was developed by Oleg I. Bydanov. During registration, the database is populated with information on medical history, histological diagnosis, disease stage, treatment administered both at the place of residence and at the Center, and follow-up data, including overall and event-free survival.

Before inclusion in the study, CT data were anonymized at the Center using the anonymization tool of Horos DICOM Viewer (this software is not developed by a single company but by a community of developers and sponsors; it is based on the OsiriX project and other open-source libraries for medical imaging, including OpenJPEG, OpenGL, VTK, ITK, and DCMTK, among others (USA). After anonymization, each patient was assigned a unique identification number.

The target pathological finding in this study was a lesion with a mean diameter of at least 4 mm detected on chest CT in a patient with OS.

The study involved staff from the following institutions: Dmitry Rogachev NMRC PHOI, AIRI Institute, Sechenov University, NSMU, AIRA Labs, and RPCC DTT.

### Eligibility criteria

#### Inclusion criteria

The study included data from patients aged 13-18 years with OS, with or without pulmonary metastatic involvement, either at initial diagnosis or at disease recurrence. An additional inclusion criterion was the availability of chest CT performed during the initial disease workup or at the time of recurrence confirmation.

This age range was selected to make the patients’ anthropometric parameters – namely body size, thoracic volume, and chest circumference – as close as possible to those of adult patients, thereby minimizing potential error of the AI algorithm trained on adult data.

Pulmonary CT examinations were performed according to chest CT acquisition protocols approved by the respective medical institution. In this study, no effort was made to specify the institution where CT was performed if the examination had not been carried out at the Center.

#### Exclusion criteria

1. History of chest trauma or surgery.
2. Chest CT findings of pleural effusion, pneumothorax, the presence of foreign bodies (tracheostomy tube, drains), motion artifacts, severe thoracic deformities (kyphoscoliosis, pectus carinatum), or other pulmonary and mediastinal pathology (abscess, emphysema, pleural empyema, etc.), as well as cardiomegaly and thymomegaly.
3. Incomplete chest scanning. A complete scan range was defined as coverage from the lung apices to the diaphragmatic domes, including the costophrenic recesses, acquired during breath-hold at full inspiration.

No **non-inclusion criteria** were planned.

### Diagnostic methods

#### Investigated diagnostic method

The study was conducted using an AI algorithm developed by AIRA Labs, for which a medical device registration certificate was obtained from the Federal Service for Surveillance in Healthcare (Roszdravnadzor): the software for CT analysis using AI technologies, Intelligent Radiology Assistants, Technical Specification 58.29.32-001-44270315-2021, registration certificate No. RZN 2024/22895, developed by AIRA Labs. Independent expert evaluation performed by specialists from the RPCC DTT showed that the algorithm achieved an AUC of 0.98 for detecting suspicious focal pulmonary lesions classified as findings requiring exclusion of malignant involvement; sensitivity was 0.94, specificity 0.98, and accuracy 0.96.

The diagnostic method under investigation is a fixed version of the AI algorithm for automated detection of pulmonary nodules on chest CT images, LungNodulesIRA. The algorithm takes a volumetric chest CT study as input; prior to application of the main model, the lungs are segmented, the voxel spacing is resampled to a uniform spatial resolution, and intensity values are normalized. The main model operates in two stages: the first stage performs highly sensitive localization and segmentation of all findings with a non-zero probability of corresponding to pulmonary nodules; the second stage applies a more specific classification of the detected objects into the classes “pulmonary nodule,” “lymph node,” “calcification,” and “other.” Based on the segmentation masks, the algorithm also estimates the volume of all findings. As a result, findings classified as pulmonary nodules with a volume greater than 50 mm3 are visualized on an additional DICOM series overlaid on the original CT images. If such findings are present, smaller nodules, lymph nodes, and calcifications are also visualized for additional review by the radiologist. If the model does not detect pulmonary nodules with a volume greater than 50 mm3, no additional findings are visualized on the DICOM series, thereby helping to avoid unnecessary cognitive load for the physician.

The model was trained using both public and internal annotated CT datasets. The segmentation component used the LIDC-IDRI dataset, which includes 1,018 CT examinations with pulmonary nodule annotations provided by multiple radiologists, and the NSCLC-Radiomics dataset, which includes 400 CT examinations with manual annotation of tumor lesions. In addition, an internal dataset of 1,243 CT examinations acquired in 2021-2022 was used; expert annotation classified algorithm-detected objects into the categories “pulmonary nodule,” “lymph node,” “calcification,” and “other.” During training of the two-stage model, the weights of the pre-trained sensitive segmentation model were used; during training of the object classification component, the segmentation backbone weights were frozen, while the parameters of the additional branch and classification head were optimized on expert-annotated candidates. The software implementation is based on the PyTorch framework; the full training cycle takes approximately 5 days on a single NVIDIA A40 GPU.

Method performance was evaluated on a held-out set of 1,247 CT examinations reflecting the clinical CT workflow. For each examination, data were available on the presence and number of pulmonary nodules, their location, and the size of the largest nodule; these data were derived from physician reports and expert review of cases with discrepancies between physician assessment and algorithmic predictions. Algorithm performance was assessed at the patient level: the unit of analysis was an individual CT examination, for which the reference status of pulmonary nodule presence and the numerical probability score produced by the model were compared. For this setting, the ROC AUC was calculated, reflecting the model’s ability to assign higher scores to examinations with pulmonary nodules than to examinations without them; for the investigated version of the model, the ROC AUC was 0.94.

In the present study, we evaluated the ability of the AI algorithm to detect intrapulmonary focal lesions potentially corresponding to pulmonary metastases when the lesion size was at least 4 mm and/or the volume was at least 50 mm3. This threshold was selected not as a universal criterion for metastasis, but as a reproducible criterion for a detectable object, consistent with the technical characteristics of the algorithm and ensuring comparability between AI assessment and expert annotation.

#### Reference Diagnostic Method

Each native chest CT was independently reviewed by a radiologist from the Center (a collective term encompassing three physicians with 7, 10, and 17 years of experience in radiation diagnostics in pediatric and adult oncology) and by two experts from AIRA Labs, CT specialists with 7 and 8 years of radiology experience, respectively. In all cases, the Center physician used DICOM viewing software (until 2016: OsiriX DICOM Viewer, developed by a community of developers and sponsors, USA; from 2016: Horos DICOM Viewer and RadiAnt DICOM Viewer, developed by Medixant, Poland). Experts from AIRA Labs and the expert from RPCC DTT evaluated the CT scans using RadiAnt DICOM Viewer (Medixant, Poland).

Each specialist searched for foci on CT images of the lungs, including calcified foci, with a mean size ≥4 mm (mean size defined as the arithmetic mean of the long and short diameters) and volume ≥50 mm^3^. The thresholds of pulmonary nodules ≥4 mm / ≥50 mm^3^were established as the minimum cutoff based on screening programs for detecting MN in the lungs [17], where this criterion was used as a reproducible criterion for a detectable intrapulmonary nodule rather than as an independent criterion for metastatic involvement.

The radiologist’s opinion from the Center was the conclusion formed during routine clinical practice from January 2012 to March 2025 and documented electronically in the medical information system. AIRA Labs experts issued electronic conclusions describing each chest CT from March to December 2025. Tomogram evaluation results were available to the principal investigator for comparison and assessment of agreement between conclusions. During the principal investigator’s comparison, if at least one focus of the target size/volume was present in the conclusions of both specialists (Center and AIRA Labs), the study was designated as “with pathology.” If neither conclusion indicated any target focus, the study was designated as “normal.” Documentation of studies “with pathology” and “normal” was performed manually by the principal investigator in a table titled “Final Study Results” accessible to co-investigators (corresponding to Table 1 in Appendix 1 of this study). In cases of disagreement between conclusions, the principal investigator classified the case as “difficult” and referred it to an expert from RPCC DTT with 29 years of radiology experience for a final decision. The expected outcome from the expert was a third opinion following binary distribution (study “with pathology” / “normal”), concordant with either the Center radiologist’s opinion or the AIRA Labs experts’ opinion.

Subsequently, all chest CTs in the dataset were analyzed using the AI algorithm. Schematically, the study workflow is presented in Figure 1:

**Figure 1.**
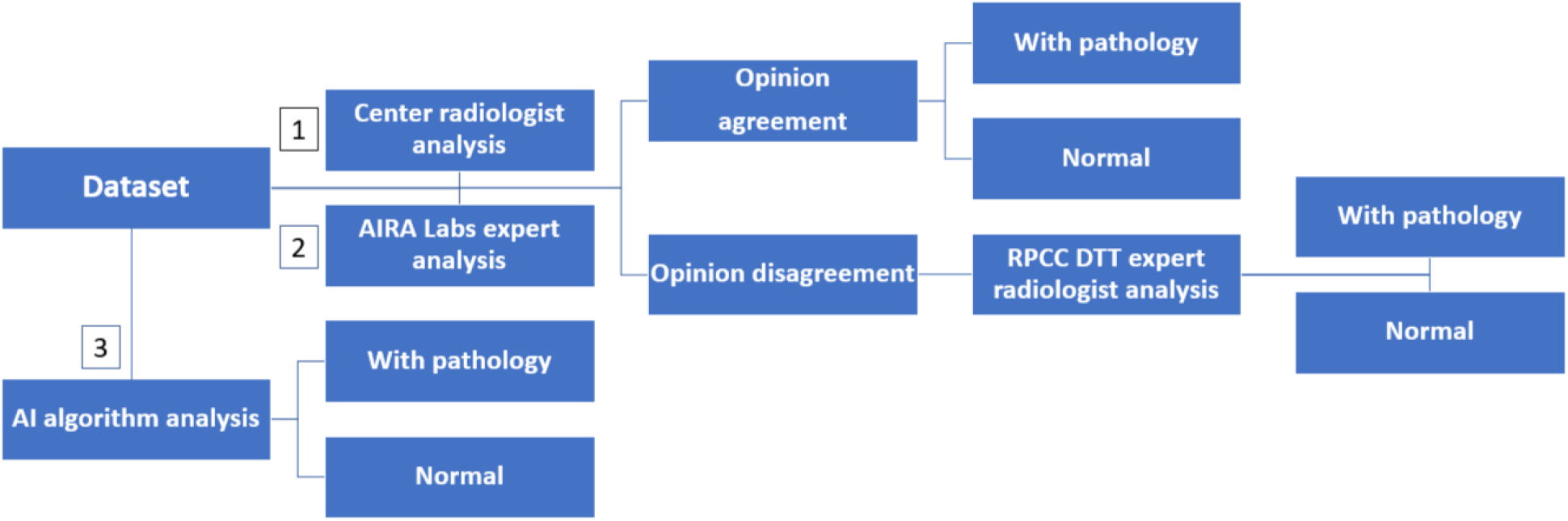
Study workflow (numbers indicate the chronological sequence of dataset analysis stages)

#### Diagnostic Accuracy Metrics

The number of false positives (FP) and false negatives (FN), sensitivity, specificity, accuracy, positive predictive value (PPV), negative predictive value (NPV), and F1-score were evaluated.

An FP was defined as a result where the AI algorithm detected a target pathological finding on tomograms that, according to two specialists (three in difficult cases), showed no radiological signs of focal lung involvement.

An FN was defined as a result where the AI algorithm failed to detect a target pathological finding on tomograms that, according to two specialists (three in difficult cases), showed radiological signs of focal lung involvement.

Sensitivity was calculated as the proportion of true positive (TP) results (correctly identified CTs with lung foci) among all “with pathology” cases (TP/(TP+FN)).

Specificity was calculated as the proportion of true negative (TN) results (correctly identified CTs without lung foci) among all “normal” cases (TN/(TN+FP)).

Accuracy was calculated as the overall proportion of correct results (both TP and TN) among all CTs (both “with pathology” and “normal”), (TP+TN)/(TP+TN+FP+FN).

PPV was calculated as the proportion of CTs with foci among cases where AI detected lung foci (TP/(TP+FP)).

NPV was calculated as the proportion of CTs without foci among cases where AI did not detect lung foci (TN/(TN+FN)).

The F1-score was calculated as the harmonic mean of sensitivity and PPV and was used to evaluate the balance between these two metrics.

For each metric, the 95% confidence interval (95% CI) was calculated.

AUC was calculated from the ROC curve constructed based on the binary classification output of the algorithm to assess the AI algorithm’s ability to distinguish cases with foci from those without (“with pathology” vs. “normal”).

The algorithm’s performance was considered satisfactory when the area under the characteristic (ROC) curve (AUC) was ≥0.81 [18].

**Sensitivity analysis** was not preplanned.

### Statistical Procedures

#### Planned Sample Size

The minimum sample size was determined by a priori statistical power analysis for evaluating the AI algorithm’s sensitivity in detecting pathological lung nodules. The minimum sample size n for estimating sensitivity was calculated using the formula for a binomial proportion with a specified CI width: n=(Z (1-α/2)^2 “ “ p(1-p))/d^2, where p is the expected sensitivity of the algorithm, d is the maximum allowable half-width of the CI, and Z_(1-α/2) is the quantile of the standard normal distribution. Calculations assumed an anticipated algorithm sensitivity of at least 0.80, a significance level α=0.05, and a desired 95% CI width of no more than ±0.08. Calculations were performed in R (packages pwr, pROC) and GraphPad Prism 10.2.2.397 (GraphPad Software Inc., USA).

Based on the calculation, the sample size was n=59. Since the study included approximately equal numbers of cases (50% patients with metastases and 50% without metastases), the total sample size was N=2×59=118. Thus, to obtain a reliable estimate of sensitivity, specificity, and AUC ROC with a CI of ±0.10 at a significance level of 0.05, the minimum required sample was 118 CT studies (conditionally: 59 with metastases and 59 without).

Due to potential data loss and the need to increase statistical power, the sample size was increased to approximately 200 studies (∼100 with metastases, ∼100 without).

*Rationale for increasing the sample size:*

1. Compensation for possible data exclusions: some images may be removed due to low quality, artifacts, or annotation errors.
2. Improved accuracy of AUC ROC estimation: larger samples enhance ROC analysis stability.
3. Guaranteed achievement of the ≥0.81 threshold for sensitivity and specificity: accounting for potential fluctuations in real data reduces the risk of random deviations.
4. Universal approach to diagnostic studies: according to recommendations, sample sizes in diagnostic accuracy studies are increased by 20% to compensate for methodological losses.

Thus, the final sample size was approximately 200 CT studies (∼100 with metastases and ∼100 without), which would allow reliable evaluation of the AI model’s diagnostic accuracy.

#### Statistical Methods

Data analysis was performed using RStudio 9.5 Build 191742 (Posit PBC, USA) with the irr and pROC libraries. GraphPad Prism 10.2.2.397 (GraphPad Software, Inc., USA) was used for statistical sample size estimation. For accuracy, sensitivity, specificity, PPV, and NPV, 95% CIs were calculated using the Wilson method; for the F1-score, the bootstrap method was used. For paired comparisons of accuracy, sensitivity, and specificity, McNemar’s exact test was applied. Differences were considered statistically significant at p<0.05.

## RESULTS

### Study Participants

Formation of the study participant sample is shown in Figure 2.

**Figure 2.**
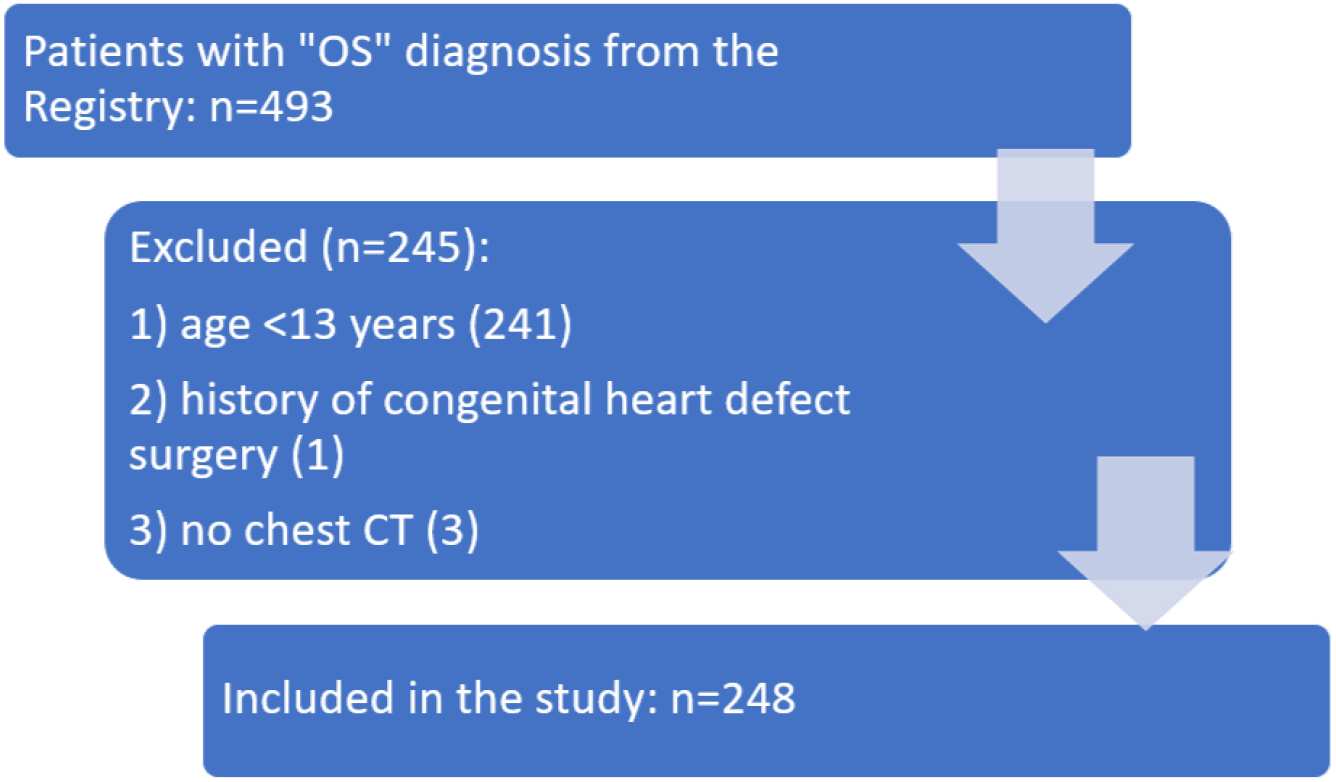
Study sample formation. Note: The period of patient registration in the Registry from January 2012 to March 2025 was considered (March 25, 2025 – the date of study synopsis approval by the Center’s professional expert commission).

The final sample consisted of 248 patients (62.5% male; mean age 15.29±1.37 years). Results of chest CT analysis for all 248 patients by two specialists, and in difficult cases by three, are presented in Table 1 (see Appendix 1).

### Results of Diagnostic Tests

An analysis of the Center physician’s integral clinical decision, reflecting the work of all specialists involved in describing CT studies issued during routine prospective evaluation of 248 tomograms, was performed: a target finding was identified in 104 cases, marked “pathology present”; in 144 cases, no target finding was identified, marked “normal” (Table 1, column 2).

At the dataset evaluation stage by AIRA Labs experts, 120 cases with target pathology (“pathology present”) and 128 without (“normal”) were identified (Table 1, column 4).

When comparing results with the Center physician’s opinion (Table 1, columns 2 and 4), agreement was observed in 212 CTs (85.5%): in 94 cases, the Center physician and AIRA Labs experts unanimously agreed on the presence of target pathology, and in 118 cases on its absence. Disagreement was observed in 36 cases (14.5%). These 36 cases were classified as “difficult” and evaluated by an expert radiologist from RPCC DTT (Table 1, column 5). There were no cases where the RPCC DTT expert could not reach a conclusion. No disagreement between the third expert’s opinion and either the first or second was recorded. No uncertain conclusions outside the binary distribution “pathology present” / “normal” were given by any expert in any case.

Based on the two (in difficult cases, three) evaluations by the Center physician, AIRA Labs experts, and the RPCC DTT expert radiologist, a final medical opinion was finalized for all 248 CTs (Table 1, column 6): target pathology was confirmed in 128 cases (51.6%), and absence of target pathology in 120 cases (48.4%).

Based on dataset evaluation using the AI algorithm, 99 tomograms with pathology (“pathology present”) and 149 without (“normal”) were identified (Table 1, column 3). AI algorithm results were not used in forming the final expert reference.

When comparing the consolidated medical opinion (column 6) with the AI algorithm evaluation result (column 3), the following data were obtained: in 5 (2.0%) cases, the AI algorithm showed FP, in 34 (13.7%) – FN, and in 209 (84.3%) – correct result (both TP and TN). The distribution of FP results (3 cases – intrapulmonary lymph nodes (60%), 2 – vascular bifurcations (40%)) reflects the model’s tendency to label certain normal anatomical structures and nonspecific changes as potential foci requiring visual verification by a physician.

Of 128 CTs “with pathology,” the AI algorithm correctly classified 94 studies (73.4%); of 120 CTs “normal,” it correctly classified 115 studies (95.8%).

When comparing the Center physician’s CT evaluation results with the consolidated medical opinion without considering AI algorithm results, it was found that target pathology (focus ≥4 mm) was missed by the Center physician in 25 of 128 TP cases (FN count was 19.5%). Of these 25 cases, the AI algorithm detected it in 11 (44%). In 4 of 11 cases, the focus was not detected at all by the Center physician (patients #60, 101, 107, 248). Notably, in all these cases, the pathological focus was located in the basal lung segments (Figure 3).

**Figure 3.**
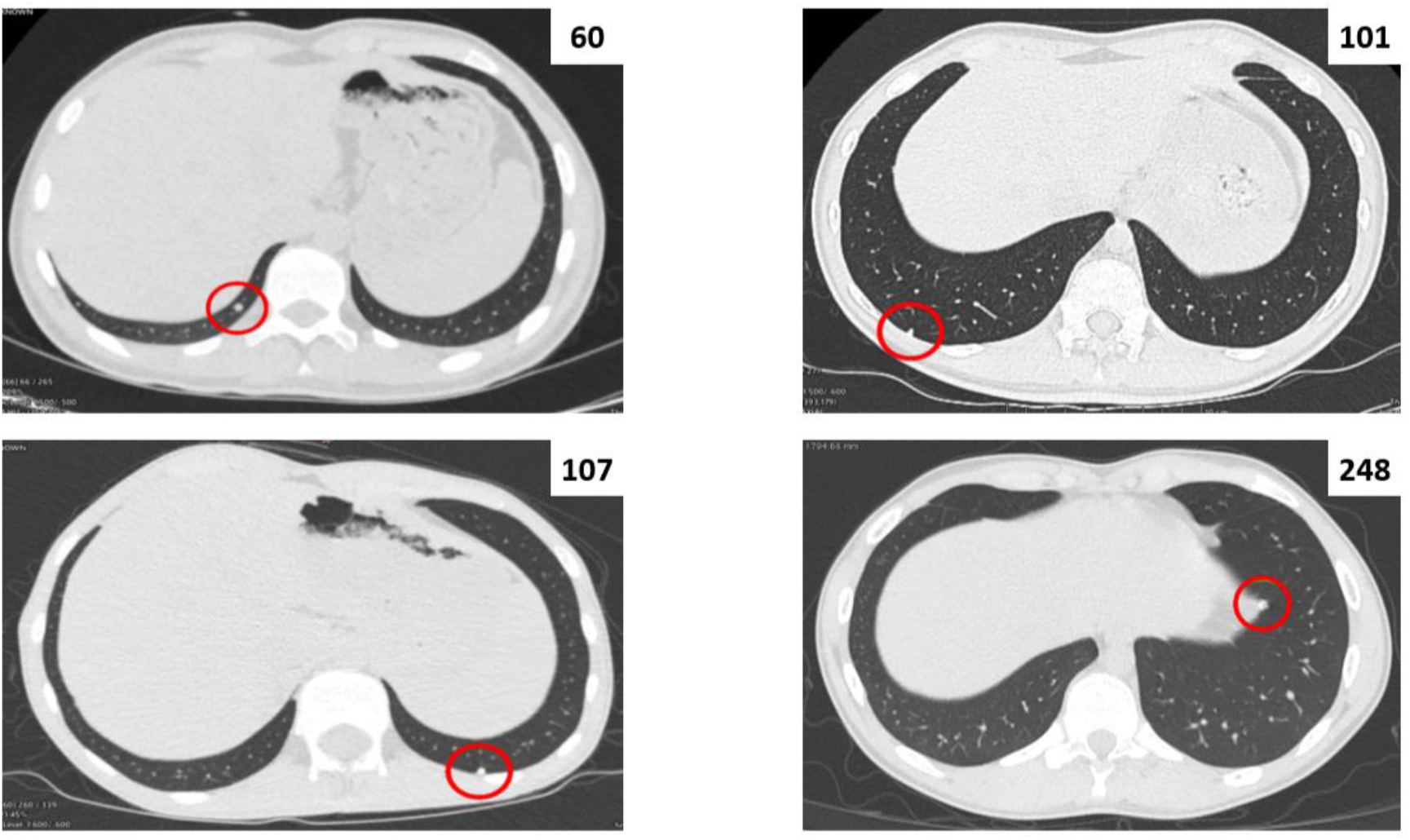
Chest CT images of patients with target pathological findings in the basal lung segments.

In 4 cases (patients #68, 146, 230, 245), findings were detected but described not as pathological but as a zone of consolidation/fibrosis (#68) or as an intrapulmonary lymph node (#146, 230, 245). In 3 cases (#159, 183, 246), the Center physician performed incorrect focus measurements, unintentionally reducing its mean size, resulting in these cases being classified as “normal.”

Table 2 presents the direct results of dataset evaluation by the Center physician and AI algorithm, as well as the result of the hypothetical strategy “Center physician and AI algorithm”:

**Table 2.** Dataset evaluation results.

| Method | TP | FP | FN | TN |
| --- | --- | --- | --- | --- |
| Center Physician | 103 | 1 | 25 | 119 |
| AI algorithm | 94 | 5 | 34 | 115 |
| Center physician and AI algorithm | 114 | 6 | 14 | 114 |

Table 3 presents the main diagnostic metrics with 95% CIs for the Center physician and AI algorithm, as well as the “Center physician and AI algorithm” strategy:

**Table 3.**
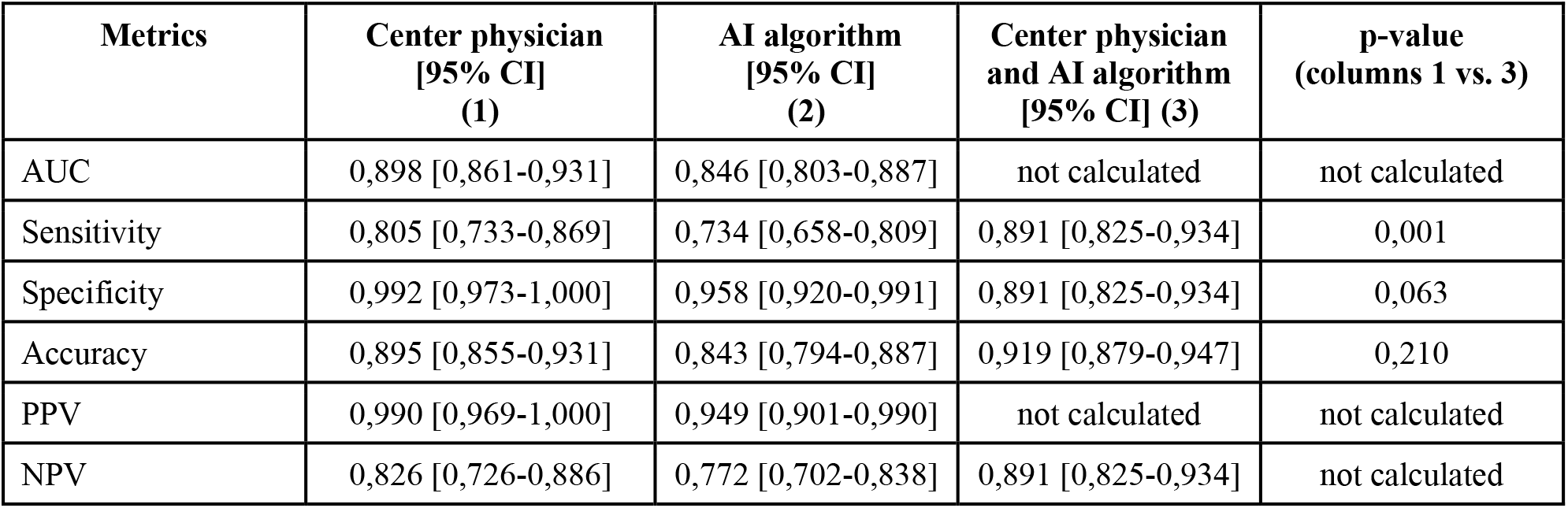
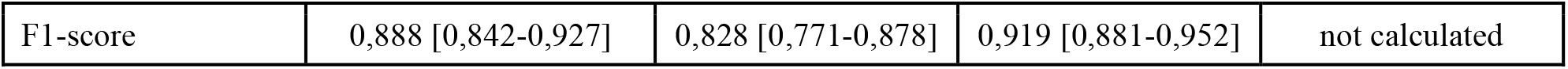
Results of statistical data analysis.

The AI algorithm showed lower accuracy than the Center physician: 0.843 vs. 0.895, p=0.047. Differences in sensitivity and specificity also did not reach statistical significance: 0.734 vs. 0.805, p=0.150 and 0.958 vs. 0.992, p=0.219, respectively.

## DISCUSSION

Among Russian studies on the declared topic published at the time of manuscript preparation, none were found.

### Summary

The point estimate of diagnostic accuracy of the applied AI algorithm was approximately 0.84, with the lower bound of the 95% CI at 0.79, indicating potential clinical value of the algorithm while simultaneously reflecting remaining statistical uncertainty and the need for confirmation in a larger and more representative sample.

### Discussion of Main Study Result

The purpose of this study was to evaluate the quality of lung focus diagnosis in adolescents with OS using AI. The results demonstrate lower diagnostic accuracy (0.84) of the applied AI algorithm in pediatric patient data compared to physician diagnostic accuracy (0.90) and significantly lower accuracy compared to adult patients (0.96). Results align with conclusions from a limited number of previous studies on this topic.

In the publication by Ni et al. [10], dedicated to detection and evaluation of pathological lung foci in OS patients using a deep convolutional neural network (DCNN), the AUC for DCNN was 0.795, and for the manual method was 0.687. The AI study targeted detection of calcified, solid, part-solid nodules, and ground-glass nodules on 675 chest CTs from 109 OS patients aged 9–35 years. In our study, we did not emphasize focus density characteristics, considering only size (target ≥4 mm) and volume (target ≥50 mm^3^). In contrast, Chinese colleagues detected foci from 2 mm, which likely contributed to reduced diagnostic accuracy in their study. Tomogram analysis was performed by two physicians with 5 and 8 years of radiology experience (in our case – three radiologists with 7, 10, and 17 years of experience). The following parameters were evaluated: sensitivity, specificity, AUC, and CT analysis time. DCNN showed higher sensitivity (0.923 vs. 0.908) and specificity (0.552 vs. 0.351) compared to the manual method, without reporting 95% CIs. In our study, the AI algorithm showed lower sensitivity (0.734 [95% CI 0.658–0.809]) but higher specificity (0.958 [95% CI 0.920–0.991]). CT reading time was significantly reduced with DCNN (173 sec) compared to the manual method (328 sec). This parameter was not evaluated in our study. Notably, in the Chinese colleagues’ publication, DCNN performance was compared with results from physicians with less experience (junior physicians) than in our study, which may have amplified the observed algorithm superiority effect. In contrast, our study compared against clinical evaluation at a reference center where foci are assessed by experienced specialists (senior physicians), making it more difficult to achieve a similar metrics gap and limiting direct comparability between studies.

It should also be noted that one objective of the Ni et al. study was monitoring dynamic changes in nodules ≤5 mm in diameter in the lungs using DCNN to approximately assess whether they are benign or metastatic. Thus, in 40 patients, dynamic CT image analysis detected changes in foci: increase or decrease in quantity, increase in density and diameter. According to the authors, increased nodule size and increased number of calcified and solid nodules indicate a higher probability of metastasis. In our study, CTs from 248 unique patients were evaluated; dynamic tomograms of the same patient were not included in the analysis, which precluded monitoring dynamic focus changes.

In the article by Salman et al. [11], the effectiveness of an AI tool for detecting lung nodules in adults (Syngo CT Lung Computer-Aided Detection, CAD) was evaluated when applied to chest CT assessment in children. Lung CAD is intended for adult use with mean solid nodule diameter ≥3 mm and subsolid nodules ≥5 mm. Thirty chest CTs with or without contrast from patients aged 12–18 years for various clinical indications were analyzed (of 30, 19 (63%) had MN without specified diagnosis). The authors emphasize age restrictions in patient enrollment to approximate adult sizes. A similar age-based sample restriction approach was used in our study. All chest tomograms, unlike CTs in our patients, were performed on a single scanner, and the diagnostic procedure was standardized. CT analysis was performed by two radiologists with 15 and 13 years of experience (senior physicians); Lung CAD results were compared with reference reading based on each nodule by two other radiologists with 2 and 6 years of experience (junior physicians). During the work, the number, size, location, and density (solid and subsolid) of nodules were determined. AI sensitivity and PPV were evaluated compared to consensual opinion of two radiologists depending on CT slice thickness: at 1 mm slice thickness, AI sensitivity and PPV were 39% and 62%, respectively; at 3 mm – 26% and 48%, respectively. In our study, statistical data analysis by CT slice thickness, nodule density character, and size (except for target 4 mm) was not performed. When excluding solid nodules <3 mm and subsolid nodules <5 mm from analysis, sensitivity at 1 mm slice was 68%, at 3 mm – 49%, but PPV did not change substantially (60% at 1 mm and 48% at 3 mm). Thus, regardless of slice thickness and nodule size, the adult CAD system showed low sensitivity when applied to pediatric patients. In our study, the AI algorithm showed higher sensitivity (0.734 [0.658–0.809]) despite methodological differences in focus evaluation.

The objective of Hardie et al. [12] was to evaluate diagnostic accuracy of various CAD systems (traditional FlyerScan and deep learning MONAI), developed and tested on adult patients, for detecting lung nodules on chest CTs in children and comparing their generalization ability in children and adults. Sensitivity and FP count at the nodule level for each chest CT were investigated (in our work, evaluation was not at each pathological finding level but at each CT level). The retrospective study included pediatric and adult CT datasets: the first consisted of 59 CT scans from 59 children with mean age 13.1 years (range 4–17 years) with suspected or confirmed MN; the second was the publicly available Lung Nodule Analysis (LUNA) 2016 subset 0 containing 89 anonymized scans with pre-labeled nodules. All studies were performed on a single scanner, and the CT procedure was standardized as in Salman et al. Scans were analyzed by two certified pediatric radiologists with additional training in focal lung lesion detection, with 2 and 11 years of experience (junior physicians). Radiologists identified 1355 nodules, including 288 nodules 3–30 mm and 1067 nodules 1–3 mm; findings >30 mm were omitted. Both scanning systems showed significantly lower lung nodule detection sensitivity in pediatric patients (68.4 [95% CI 65.1–73.0] and 53.1% [95% CI 46.7–58.4], respectively) compared to adult dataset data (83.9 [95% CI 79.1–88.0] and 95.5% [95% CI 90.0–98.4], respectively). In our study, AI sensitivity in adolescent CT analysis (0.734 [95% CI 0.658–0.809]) was approximately comparable but slightly higher than FlyerScan and significantly higher than MONAI; when searching for lung foci in the adult reference dataset, AI showed sensitivity of 0.94. The authors emphasize the significant (p<0.001) difference in mean nodule size in pediatric test data (5.4±3.1 mm) compared to adult dataset data (11.0±6.2 mm): smaller nodules (including 1–3 mm) predominated in pediatrics and were underrepresented in adult training samples. Additionally, the study included preschool and early school-age patients with anatomical differences from adults (smaller chest size, denser vascular network). Another point with which we agree is the differences in lung metastasis character in children compared to primary lung MN in adults. The authors believe these factors caused low sensitivity of both AI models in pediatric CT analysis. It should be noted that Salman et al. [11] and Hardie et al. [12] included data from patients without MN and with various unspecified MN, unlike Ni et al. [10] and our work, which focused exclusively on OS.

Reiterating, unlike Ni et al.’s metrics, AI sensitivity in this work was substantially lower at 0.734, and specificity was incomparably higher at 0.958.

Probable reasons for this sensitivity include: the AI algorithm was trained on adult patients and previously had not been applied to pediatric-adolescent data. In the current study, we specifically limited the minimum age to 13 years to reduce this effect, but it persists. Additionally, we evaluated at the level of each specific study (chest CT of a specific patient), while colleagues from China [10] and the USA [11, 12] quantitatively evaluated detection and omission of individual lung nodules. These fundamentally different approaches yield different error profiles and make sensitivity not fully comparable: at nodule-level evaluation, an algorithm may show very high sensitivity by “finding at least something resembling a nodule”; at study-level evaluation, any missed target finding immediately converts the case to FN. Additionally, the AI algorithm training sample differs in target finding characteristics; in particular, the most common feature of metastatic lung foci in OS is calcification [10, 19, 20], while our AI algorithm focuses on detecting soft-tissue foci. In Salman et al.’s study, none of the 109 nodules detected by radiologists were calcified, which may indirectly indicate absence of OS patients in that study [11].

Sensitivity and specificity for the Center physician were 0.805 [95% CI 0.733–0.869] and 0.992 [0.973–1.000], respectively. In our opinion, the Center physician’s sensitivity was lower than in Ni et al. (0.908) [10] because Chinese colleagues worked under a laboratory scenario for a retrospective study where experts had more time and a specific goal compared to Center physicians whose opinion was formed during prospective work in real clinical practice. Additionally, in three cases the Center physician inadvertently made measurement errors, and in four cases interpreted detected changes as intrapulmonary lymph nodes or fibrosis. Missed foci may also have been influenced by predominant localization in basal lung segments, where detection is significantly more difficult due to respiratory artifacts.

The very high specificity of the Center physician (0.992, vs. 0.351 in Ni et al. [10]) was related to a more cautious clinical approach: the physician less frequently classifies questionable structures as pathological, so there are almost no FP conclusions in their evaluation (1 case). Additionally, dataset formation in the absence of histological data may have influenced the indicator: the reference standard was based on expert consensus including the Center physician’s opinion.

The DCNN model in Ni et al.’s study identified 3087 nodules, missing 278 (nodule omission rate 8.3%); when analyzing the same images using the manual radiologist method, 2442 nodules were detected, 657 missed (21.2% nodules) [10].

In Salman et al.’s study, at 1 mm slice thickness the CAD system detected 26 FP nodules out of 70 found; at 3 mm slice thickness – 30 out of 60. Examples of such findings included scars, peripheral consolidation zones, blood vessels. The observed large number of FP results substantially impacted low AUC values [11]. In our study, the AI algorithm showed only 5 FP results, constituting 2% of all cases (vascular bifurcations and intrapulmonary lymph nodes).

Possibly, to increase sensitivity in lung focus diagnosis in children, specificity must be reduced. Such an approach would undoubtedly increase FP results requiring radiologist verification but could help mitigate the observed sensitivity reduction. In our study, adding the AI algorithm to the Center physician’s conclusion reduced specificity from 0.992 to 0.891, indicating that AI algorithm suggestions should be considered not as a final radiological diagnosis but as a reason for targeted CT re-evaluation by a physician. Such AI usage may be justified when missed findings have high clinical significance and in conditions of high radiologist workload in reference centers with limited review time.

According to Patel et al.’s 2020 study, CT interpretation error by a radiologist is approximately 3– 5% under normal conditions and increases several-fold in difficult cases and adverse conditions (e.g., overnight shifts) [21]. According to Gefter et al., the average radiologist error rate is approximately 4% in typical cases evaluating studies without pathology, and when considering only pathological findings, the error rate may reach 30% or higher [22]. Zhang et al. report similar figures [23].

For metastasis detection, the most important metric is sensitivity because the clinical cost of missing a pathological focus is higher than the cost of additional expert FP verification, and an FN result can lead to underestimation of disease stage and inappropriate treatment changes. Therefore, during the study we separately formulated and calculated a scenario where the AI algorithm is used to assist the physician as a second reader-assistant. In this scenario, the Center physician using the AI algorithm would miss the target finding in 14 of 128 TP cases instead of 25 (FN count, simply put, missed foci would decrease by 8.6% to 10.9%). Meanwhile, the Center physician would receive only 5 additional FP results requiring repeated expert analysis in real clinical practice.

The absence of statistically significant differences between accuracy, sensitivity, and specificity of AI algorithm and Center physician work does not permit using AI as an independent tool for CT evaluation, but the high metric values indicate potential value as a second reader-assistant. The DCNN proposed by Ni et al., according to the authors, can be used as an effective tool for detection and monitoring of lung nodules in OS patients [10].

When applying the AI algorithm in the “physician and assistant” scenario, a significant increase in all statistical data analysis metrics is observed, primarily a statistically significant increase in sensitivity (p<0.05), so using the AI algorithm as an assistant can and should underlie radiologist practice in solving the clinical task of lung focus detection in adolescent OS patients.

Diagnostic accuracy of the applied AI algorithm obtained in this study, equal to 0.84, exceeded the established diagnostic accuracy threshold determined by AUC of 0.81. However, when interpreting algorithm diagnostic metrics for clinical practice, it is more appropriate to focus on the lower bound of the 95% CI. In this study it was 0.79, indicating persistent statistical uncertainty and emphasizing the need for algorithm validation on a larger and more representative sample before widespread clinical implementation.

### Study Limitations

Our study has several limitations.

1. In this study, patients from the Center Registry included in the study underwent primary examination not only at Center but also in various regions of the Russian Federation. Nevertheless, low-dose chest CT was not performed in any of the 248 cases included in the analysis.
2. Limitations also include the retrospective nature and evaluation within a single institution with a relatively small number of patients. Due to OS rarity in children and adolescents and the Registry’s single-center character, forming a representative and standardized dataset for AI algorithm training and validation appears difficult. In this study, to ensure greater sample homogeneity, patients under 13 years were excluded from the overall OS cohort (493), which somewhat limits result extrapolation to the entire pediatric and adolescent population with this disease. The AIRA Labs AI algorithm used in this study had not previously been applied to pediatric and adolescent patient data. Possibly, this circumstance caused reduced statistical metric values (especially AUC). In this study, a preplanned sensitivity analysis with separate subgroups by age or anthropometric indicators was not conducted due to limited overall sample size for this rare disease and potentially high statistical uncertainty. Detailed study of age, body build, and CT protocol parameter effects on AI algorithm performance requires a separate study with sufficient sample size in relevant subgroups.
3. The AI algorithm detected intrapulmonary foci potentially corresponding to metastatic involvement; metastasis status was established postfactum based not on clinical-radiological data but on planned histological examination if performed. Additionally, CTs from 248 unique patients were evaluated; dynamic tomograms of the same patient were not included in the analysis, precluding monitoring of dynamic focus changes. Furthermore, correlation analysis between CT-detected lung foci and their morphology was not a study objective.
4. The final medical opinion (final reference standard) was not fully independent, as it was formed using an arbitration scheme with primary medical evaluations: when Center physician and AIRA Labs expert conclusions matched, this agreement was accepted as the final verdict; in case of disagreement, a referee expert with 29 years of experience was involved. Therefore, metrics of Center physician work relative to the final reference obtained in this study reflect the degree of correspondence to the arbitration expert decision, not absolute diagnostic accuracy relative to a fully independent external standard. AI algorithm results were not used in forming the final expert reference, reducing incorporation bias risk for AI algorithm evaluation. Additionally, the “Center physician and AI algorithm” scenario is a retrospective model of AI algorithm use as a second reader- assistant and requires confirmation in prospective clinical workflow.
5. Statistical criterion power evaluation was performed post hoc based on actually obtained data, reflecting achieved but not a priori specified statistical power.

## CONCLUSION

The results obtained in this study indicate the practical value of applying an AI algorithm for the specific clinical task of detecting pathological lung foci in OS patients. Using AI improves diagnostic accuracy of lung foci, thereby accelerating the process of making correct clinical decisions for each individual patient. To improve focus evaluation accuracy and differential diagnosis in lungs, further research in this knowledge area is necessary with emphasis on developing specialized pediatric systems and creating specific training datasets for pediatric and adolescent patients.

## Data Availability

All data produced in the present work are contained in the manuscript

