## Appendix 1. Table 1. for "Assessment of the accuracy of lung lesions diagnosis in adolescents with osteosarcoma using artificial intelligence"

| No. | Center physician's opinion | AI algorithm | Opinion of AIRA Labs experts | Expert radiologist’s opinion from RPCC DTT | Consolidated medical opinion | AI algorithm output |
| --- | --- | --- | --- | --- | --- | --- |
| 1 | normal | normal | with pathology | with pathology | with pathology | FN |
| 2 | normal | with pathology | normal | - | normal | FP |
| 3 | normal | normal | normal | - | normal | correct result (both TP and TN) |
| 4 | with pathology | with pathology | with pathology | - | with pathology | correct result (both TP and TN) |
| 5 | with pathology | with pathology | with pathology | - | with pathology | correct result (both TP and TN) |
| 6 | normal | normal | normal | - | normal | correct result (both TP and TN) |
| 7 | with pathology | with pathology | with pathology | - | with pathology | correct result (both TP and TN) |
| 8 | normal | normal | normal | - | normal | correct result (both TP and TN) |
| 9 | with pathology | with pathology | with pathology | - | with pathology | correct result (both TP and TN) |
| 10 | with pathology | with pathology | with pathology | - | with pathology | correct result (both TP and TN) |
| 11 | normal | normal | normal | - | normal | correct result (both TP and TN) |
| 12 | normal | normal | normal | - | normal | correct result (both TP and TN) |
| 13 | normal | normal | normal | - | normal | correct result (both TP and TN) |
| 14 | with pathology | with pathology | with pathology | - | with pathology | correct result (both TP and TN) |
| 15 | with pathology | with pathology | normal | with pathology | with pathology | correct result (both TP and TN) |
| 16 | with pathology | with pathology | with pathology | - | with pathology | correct result (both TP and TN) |
| 17 | with pathology | with pathology | with pathology | - | with pathology | correct result (both TP and TN) |
| 18 | normal | normal | normal | - | normal | correct result (both TP and TN) |
| 19 | normal | normal | normal | - | normal | correct result (both TP and TN) |
| 20 | normal | normal | normal | - | normal | correct result (both TP and TN) |
| 21 | with pathology | with pathology | with pathology | - | with pathology | correct result (both TP and TN) |
| 22 | with pathology | normal | with pathology | - | with pathology | FN |
| 23 | with pathology | normal | with pathology | - | with pathology | FN |
| 24 | with pathology | normal | with pathology | - | with pathology | FN |
| 25 | with pathology | normal | normal | with pathology | with pathology | FN |
| 26 | normal | normal | normal | - | normal | correct result (both TP and TN) |
| 27 | normal | normal | with pathology | with pathology | with pathology | FN |
| 28 | with pathology | normal | normal | with pathology | with pathology | FN |
| 29 | with pathology | normal | with pathology | - | with pathology | FN |
| 30 | with pathology | with pathology | with pathology | - | with pathology | correct result (both TP and TN) |
| 31 | with pathology | normal | with pathology | - | with pathology | FN |
| 32 | with pathology | with pathology | with pathology | - | with pathology | correct result (both TP and TN) |
| 33 | with pathology | with pathology | with pathology | - | with pathology | correct result (both TP and TN) |
| 34 | with pathology | normal | with pathology | - | with pathology | FN |
| 35 | with pathology | with pathology | with pathology | - | with pathology | correct result (both TP and TN) |
| 36 | with pathology | with pathology | with pathology | - | with pathology | correct result (both TP and TN) |
| 37 | with pathology | with pathology | with pathology | - | with pathology | correct result (both TP and TN) |
| 38 | with pathology | with pathology | with pathology | - | with pathology | correct result (both TP and TN) |
| 39 | with pathology | normal | with pathology | - | with pathology | FN |
| 40 | normal | normal | normal | - | normal | correct result (both TP and TN) |
| 41 | with pathology | with pathology | with pathology | - | with pathology | correct result (both TP and TN) |
| 42 | with pathology | with pathology | with pathology | - | with pathology | correct result (both TP and TN) |
| 43 | with pathology | with pathology | with pathology | - | with pathology | correct result (both TP and TN) |
| 44 | with pathology | with pathology | with pathology | - | with pathology | correct result (both TP and TN) |
| 45 | with pathology | with pathology | with pathology | - | with pathology | correct result (both TP and TN) |
| 46 | with pathology | with pathology | with pathology | - | with pathology | correct result (both TP and TN) |
| 47 | normal | normal | normal | - | normal | correct result (both TP and TN) |
| 48 | with pathology | with pathology | with pathology | - | with pathology | correct result (both TP and TN) |
| 49 | with pathology | normal | normal | normal | normal | correct result (both TP and TN) |
| 50 | with pathology | with pathology | with pathology | - | with pathology | correct result (both TP and TN) |
| 51 | with pathology | with pathology | with pathology | - | with pathology | correct result (both TP and TN) |
| 52 | with pathology | with pathology | with pathology | - | with pathology | correct result (both TP and TN) |
| 53 | with pathology | with pathology | with pathology | - | with pathology | correct result (both TP and TN) |
| 54 | with pathology | with pathology | with pathology | - | with pathology | correct result (both TP and TN) |
| 55 | with pathology | with pathology | with pathology | - | with pathology | correct result (both TP and TN) |
| 56 | with pathology | with pathology | with pathology | - | with pathology | correct result (both TP and TN) |
| 57 | with pathology | with pathology | with pathology | - | with pathology | correct result (both TP and TN) |
| 58 | with pathology | with pathology | with pathology | - | with pathology | correct result (both TP and TN) |
| 59 | with pathology | with pathology | with pathology | - | with pathology | correct result (both TP and TN) |
| 60 | normal | with pathology | with pathology | with pathology | with pathology | correct result (both TP and TN) |
| 61 | normal | normal | normal | - | normal | correct result (both TP and TN) |
| 62 | with pathology | with pathology | with pathology | - | with pathology | correct result (both TP and TN) |
| 63 | with pathology | with pathology | with pathology | - | with pathology | correct result (both TP and TN) |
| 64 | with pathology | with pathology | with pathology | - | with pathology | correct result (both TP and TN) |
| 65 | with pathology | with pathology | with pathology | - | with pathology | correct result (both TP and TN) |
| 66 | normal | normal | normal | - | normal | correct result (both TP and TN) |
| 67 | with pathology | with pathology | with pathology | - | with pathology | correct result (both TP and TN) |
| 68 | normal | with pathology | with pathology | with pathology | with pathology | correct result (both TP and TN) |
| 69 | with pathology | with pathology | with pathology | - | with pathology | correct result (both TP and TN) |
| 70 | normal | normal | normal | - | normal | correct result (both TP and TN) |
| 71 | with pathology | with pathology | with pathology | - | with pathology | correct result (both TP and TN) |
| 72 | with pathology | with pathology | with pathology | - | with pathology | correct result (both TP and TN) |
| 73 | with pathology | with pathology | with pathology | - | with pathology | correct result (both TP and TN) |
| 74 | with pathology | with pathology | with pathology | - | with pathology | correct result (both TP and TN) |
| 75 | with pathology | with pathology | with pathology | - | with pathology | correct result (both TP and TN) |
| 76 | with pathology | with pathology | with pathology | - | with pathology | correct result (both TP and TN) |
| 77 | with pathology | with pathology | with pathology | - | with pathology | correct result (both TP and TN) |
| 78 | with pathology | with pathology | with pathology | - | with pathology | correct result (both TP and TN) |
| 79 | normal | normal | with pathology | with pathology | with pathology | FN |
| 80 | with pathology | with pathology | with pathology | - | with pathology | correct result (both TP and TN) |
| 81 | normal | normal | normal | - | normal | correct result (both TP and TN) |
| 82 | with pathology | with pathology | with pathology | - | with pathology | correct result (both TP and TN) |
| 83 | with pathology | with pathology | with pathology | - | with pathology | correct result (both TP and TN) |
| 84 | with pathology | normal | normal | with pathology | with pathology | FN |
| 85 | with pathology | with pathology | with pathology | - | with pathology | correct result (both TP and TN) |
| 86 | with pathology | with pathology | with pathology | - | with pathology | correct result (both TP and TN) |
| 87 | normal | normal | normal | - | normal | correct result (both TP and TN) |
| 88 | with pathology | with pathology | with pathology | - | with pathology | correct result (both TP and TN) |
| 89 | with pathology | with pathology | with pathology | - | with pathology | correct result (both TP and TN) |
| 90 | with pathology | with pathology | with pathology | - | with pathology | correct result (both TP and TN) |
| 91 | with pathology | with pathology | with pathology | - | with pathology | correct result (both TP and TN) |
| 92 | with pathology | with pathology | with pathology | - | with pathology | correct result (both TP and TN) |
| 93 | normal | normal | normal | - | normal | correct result (both TP and TN) |
| 94 | with pathology | with pathology | with pathology | - | with pathology | correct result (both TP and TN) |
| 95 | with pathology | with pathology | with pathology | - | with pathology | correct result (both TP and TN) |
| 96 | normal | normal | normal | - | normal | correct result (both TP and TN) |
| 97 | with pathology | normal | with pathology | - | with pathology | FN |
| 98 | with pathology | normal | normal | with pathology | with pathology | FN |
| 99 | normal | normal | normal | - | normal | correct result (both TP and TN) |
| 100 | normal | normal | with pathology | with pathology | with pathology | FN |
| 101 | normal | with pathology | with pathology | with pathology | with pathology | correct result (both TP and TN) |
| 102 | normal | normal | normal | - | normal | correct result (both TP and TN) |
| 103 | with pathology | with pathology | with pathology | - | with pathology | correct result (both TP and TN) |
| 104 | with pathology | with pathology | with pathology | - | with pathology | correct result (both TP and TN) |
| 105 | with pathology | normal | normal | with pathology | with pathology | FN |
| 106 | with pathology | with pathology | with pathology | - | with pathology | correct result (both TP and TN) |
| 107 | normal | with pathology | with pathology | with pathology | with pathology | correct result (both TP and TN) |
| 108 | with pathology | with pathology | with pathology | - | with pathology | correct result (both TP and TN) |
| 109 | with pathology | with pathology | with pathology | - | with pathology | correct result (both TP and TN) |
| 110 | with pathology | with pathology | with pathology | - | with pathology | correct result (both TP and TN) |
| 111 | with pathology | with pathology | with pathology | - | with pathology | correct result (both TP and TN) |
| 112 | with pathology | normal | with pathology | - | with pathology | FN |
| 113 | with pathology | with pathology | with pathology | - | with pathology | correct result (both TP and TN) |
| 114 | with pathology | with pathology | with pathology | - | with pathology | correct result (both TP and TN) |
| 115 | with pathology | with pathology | with pathology | - | with pathology | correct result (both TP and TN) |
| 116 | with pathology | with pathology | normal | with pathology | with pathology | correct result (both TP and TN) |
| 117 | with pathology | with pathology | with pathology | - | with pathology | correct result (both TP and TN) |
| 118 | normal | normal | with pathology | with pathology | with pathology | FN |
| 119 | with pathology | with pathology | with pathology | - | with pathology | correct result (both TP and TN) |
| 120 | normal | normal | with pathology | with pathology | with pathology | FN |
| 121 | with pathology | with pathology | with pathology | - | with pathology | correct result (both TP and TN) |
| 122 | with pathology | with pathology | with pathology | - | with pathology | correct result (both TP and TN) |
| 123 | normal | normal | normal | - | normal | correct result (both TP and TN) |
| 124 | with pathology | with pathology | with pathology | - | with pathology | correct result (both TP and TN) |
| 125 | normal | normal | normal | - | normal | correct result (both TP and TN) |
| 126 | normal | normal | normal | - | normal | correct result (both TP and TN) |
| 127 | normal | normal | normal | - | normal | correct result (both TP and TN) |
| 128 | normal | normal | normal | - | normal | correct result (both TP and TN) |
| 129 | normal | normal | normal | - | normal | correct result (both TP and TN) |
| 130 | normal | normal | normal | - | normal | correct result (both TP and TN) |
| 131 | normal | normal | normal | - | normal | correct result (both TP and TN) |
| 132 | normal | normal | normal | - | normal | correct result (both TP and TN) |
| 133 | normal | normal | normal | - | normal | correct result (both TP and TN) |
| 134 | normal | normal | normal | - | normal | correct result (both TP and TN) |
| 135 | normal | normal | normal | - | normal | correct result (both TP and TN) |
| 136 | normal | normal | normal | - | normal | correct result (both TP and TN) |
| 137 | normal | normal | normal | - | normal | correct result (both TP and TN) |
| 138 | normal | normal | normal | - | normal | correct result (both TP and TN) |
| 139 | normal | normal | normal | - | normal | correct result (both TP and TN) |
| 140 | normal | normal | with pathology | with pathology | with pathology | FN |
| 141 | normal | normal | normal | - | normal | correct result (both TP and TN) |
| 142 | normal | normal | normal | - | normal | correct result (both TP and TN) |
| 143 | with pathology | normal | normal | with pathology | with pathology | FN |
| 144 | normal | normal | normal | - | normal | correct result (both TP and TN) |
| 145 | normal | normal | with pathology | with pathology | with pathology | FN |
| 146 | normal | with pathology | with pathology | with pathology | with pathology | correct result (both TP and TN) |
| 147 | normal | normal | normal | - | normal | correct result (both TP and TN) |
| 148 | normal | normal | normal | - | normal | correct result (both TP and TN) |
| 149 | normal | normal | normal | - | normal | correct result (both TP and TN) |
| 150 | normal | normal | normal | - | normal | correct result (both TP and TN) |
| 151 | normal | normal | with pathology | with pathology | with pathology | FN |
| 152 | normal | normal | normal | - | normal | correct result (both TP and TN) |
| 153 | normal | normal | normal | - | normal | correct result (both TP and TN) |
| 154 | normal | normal | normal | - | normal | correct result (both TP and TN) |
| 155 | with pathology | normal | normal | with pathology | with pathology | FN |
| 156 | normal | normal | normal | - | normal | correct result (both TP and TN) |
| 157 | with pathology | with pathology | with pathology | - | with pathology | correct result (both TP and TN) |
| 158 | normal | normal | normal | - | normal | correct result (both TP and TN) |
| 159 | normal | with pathology | with pathology | with pathology | with pathology | correct result (both TP and TN) |
| 160 | normal | normal | normal | - | normal | correct result (both TP and TN) |
| 161 | normal | normal | normal | - | normal | correct result (both TP and TN) |
| 162 | normal | normal | normal | - | normal | correct result (both TP and TN) |
| 163 | normal | normal | normal | - | normal | correct result (both TP and TN) |
| 164 | normal | normal | normal | - | normal | correct result (both TP and TN) |
| 165 | normal | normal | normal | - | normal | correct result (both TP and TN) |
| 166 | normal | normal | normal | - | normal | correct result (both TP and TN) |
| 167 | normal | normal | normal | - | normal | correct result (both TP and TN) |
| 168 | with pathology | normal | with pathology | - | with pathology | FN |
| 169 | normal | normal | normal | - | normal | correct result (both TP and TN) |
| 170 | normal | normal | normal | - | normal | correct result (both TP and TN) |
| 171 | normal | normal | normal | - | normal | correct result (both TP and TN) |
| 172 | normal | normal | normal | - | normal | correct result (both TP and TN) |
| 173 | normal | normal | normal | - | normal | correct result (both TP and TN) |
| 174 | normal | normal | normal | - | normal | correct result (both TP and TN) |
| 175 | normal | normal | normal | - | normal | correct result (both TP and TN) |
| 176 | normal | normal | normal | - | normal | correct result (both TP and TN) |
| 177 | normal | normal | with pathology | normal | normal | correct result (both TP and TN) |
| 178 | with pathology | normal | with pathology | - | with pathology | FN |
| 179 | normal | normal | normal | - | normal | correct result (both TP and TN) |
| 180 | normal | normal | normal | - | normal | correct result (both TP and TN) |
| 181 | normal | normal | normal | - | normal | correct result (both TP and TN) |
| 182 | normal | normal | normal | - | normal | correct result (both TP and TN) |
| 183 | normal | with pathology | with pathology | with pathology | with pathology | correct result (both TP and TN) |
| 184 | normal | normal | normal | - | normal | correct result (both TP and TN) |
| 185 | normal | normal | normal | - | normal | correct result (both TP and TN) |
| 186 | normal | normal | normal | - | normal | correct result (both TP and TN) |
| 187 | with pathology | with pathology | with pathology | - | with pathology | correct result (both TP and TN) |
| 188 | normal | normal | normal | - | normal | correct result (both TP and TN) |
| 189 | normal | with pathology | normal | - | normal | FP |
| 190 | normal | normal | normal | - | normal | correct result (both TP and TN) |
| 191 | normal | normal | normal | - | normal | correct result (both TP and TN) |
| 192 | normal | with pathology | normal | - | normal | FP |
| 193 | normal | normal | normal | - | normal | correct result (both TP and TN) |
| 194 | normal | normal | normal | - | normal | correct result (both TP and TN) |
| 195 | normal | normal | with pathology | with pathology | with pathology | FN |
| 196 | normal | normal | normal | - | normal | correct result (both TP and TN) |
| 197 | with pathology | with pathology | with pathology | - | with pathology | correct result (both TP and TN) |
| 198 | normal | normal | normal | - | normal | correct result (both TP and TN) |
| 199 | normal | normal | normal | - | normal | correct result (both TP and TN) |
| 200 | normal | normal | normal | - | normal | correct result (both TP and TN) |
| 201 | with pathology | with pathology | with pathology | - | with pathology | correct result (both TP and TN) |
| 202 | normal | normal | normal | - | normal | correct result (both TP and TN) |
| 203 | normal | normal | normal | - | normal | correct result (both TP and TN) |
| 204 | normal | normal | normal | - | normal | correct result (both TP and TN) |
| 205 | normal | normal | normal | - | normal | correct result (both TP and TN) |
| 206 | normal | normal | normal | - | normal | correct result (both TP and TN) |
| 207 | normal | normal | normal | - | normal | correct result (both TP and TN) |
| 208 | normal | normal | with pathology | with pathology | with pathology | FN |
| 209 | normal | normal | normal | - | normal | correct result (both TP and TN) |
| 210 | normal | normal | normal | - | normal | correct result (both TP and TN) |
| 211 | normal | normal | with pathology | with pathology | with pathology | FN |
| 212 | normal | normal | normal | - | normal | correct result (both TP and TN) |
| 213 | normal | normal | with pathology | with pathology | with pathology | FN |
| 214 | normal | normal | normal | - | normal | correct result (both TP and TN) |
| 215 | with pathology | normal | with pathology | - | with pathology | FN |
| 216 | normal | normal | normal | - | normal | correct result (both TP and TN) |
| 217 | normal | normal | normal | - | normal | correct result (both TP and TN) |
| 218 | normal | normal | normal | - | normal | correct result (both TP and TN) |
| 219 | normal | normal | normal | - | normal | correct result (both TP and TN) |
| 220 | normal | normal | normal | - | normal | correct result (both TP and TN) |
| 221 | normal | normal | normal | - | normal | correct result (both TP and TN) |
| 222 | normal | normal | normal | - | normal | correct result (both TP and TN) |
| 223 | normal | normal | normal | - | normal | correct result (both TP and TN) |
| 224 | normal | normal | normal | - | normal | correct result (both TP and TN) |
| 225 | with pathology | normal | with pathology | - | with pathology | FN |
| 226 | normal | normal | with pathology | with pathology | with pathology | FN |
| 227 | normal | normal | normal | - | normal | correct result (both TP and TN) |
| 228 | normal | normal | normal | - | normal | correct result (both TP and TN) |
| 229 | with pathology | with pathology | with pathology | - | with pathology | correct result (both TP and TN) |
| 230 | normal | with pathology | with pathology | with pathology | with pathology | correct result (both TP and TN) |
| 231 | normal | normal | normal | - | normal | correct result (both TP and TN) |
| 232 | normal | normal | normal | - | normal | correct result (both TP and TN) |
| 233 | normal | normal | normal | - | normal | correct result (both TP and TN) |
| 234 | normal | normal | normal | - | normal | correct result (both TP and TN) |
| 235 | normal | normal | normal | - | normal | correct result (both TP and TN) |
| 236 | normal | normal | normal | - | normal | correct result (both TP and TN) |
| 237 | normal | normal | normal | - | normal | correct result (both TP and TN) |
| 238 | normal | normal | normal | - | normal | correct result (both TP and TN) |
| 239 | normal | with pathology | normal | - | normal | FP |
| 240 | with pathology | with pathology | with pathology | - | with pathology | correct result (both TP and TN) |
| 241 | normal | normal | normal | - | normal | correct result (both TP and TN) |
| 242 | normal | normal | normal | - | normal | correct result (both TP and TN) |
| 243 | normal | with pathology | normal | - | normal | FP |
| 244 | normal | normal | normal | - | normal | correct result (both TP and TN) |
| 245 | normal | with pathology | with pathology | with pathology | with pathology | correct result (both TP and TN) |
| 246 | normal | with pathology | with pathology | with pathology | with pathology | correct result (both TP and TN) |
| 247 | with pathology | with pathology | with pathology | - | with pathology | correct result (both TP and TN) |
| 248 | normal | with pathology | with pathology | with pathology | with pathology | correct result (both TP and TN) |
